# SARS-CoV-2 infection, clinical features and outcome of COVID-19 in United Kingdom nursing homes

**DOI:** 10.1101/2020.05.19.20105460

**Authors:** NSN Graham, C Junghans, R Downes, C Sendall, H Lai, A McKirdy, P Elliott, R Howard, D Wingfield, M Priestman, M Ciechonska, L Cameron, M Storch, MA Crone, PS Freemont, P Randell, R McLaren, N Lang, S Ladhani, F Sanderson, DJ Sharp

## Abstract

**Objectives:** To understand SARS-Co-V-2 infection and transmission in UK nursing homes in order to develop preventive strategies for protecting the frail elderly residents.

**Design:** An outbreak investigation.

**Setting:** 4 nursing homes affected by COVID-19 outbreaks in central London.

**Participants:** 394 residents and 70 staff in nursing homes.

**Interventions:** Two point-prevalence surveys one week apart where residents underwent SARS-CoV-2 testing and had relevant symptoms documented. Asymptomatic staff from three of the four homes were also offered SARS-CoV-2 testing.

**Main outcome measures:** All-cause mortality, and mortality attributed to COVID-19 on death certificates. Prevalence of SARS-CoV-2 infection and symptoms in residents and staff.

**Results:** Overall, 26% (95% confidence interval 22 to 31) of residents died over the two-month period. All-cause mortality increased by 203% (95% CI 70 to 336). Systematic testing identified 40% (95% CI 35 to 46) of residents, of whom 43% (95% CI 34 to 52) were asymptomatic and 18% (95% CI 11 to 24) had atypical symptoms, as well as 4% (95% CI -1 to 9) of asymptomatic staff who tested positive for SARS-CoV-2.

**Conclusions:** The SARS-CoV-2 outbreak was associated with a very high mortality rate in residents of nursing homes. Systematic testing of all residents and a representative sample of staff identified high rates of SARS-CoV-2 positivity across the four nursing homes, highlighting a potential for regular screening to prevent future outbreaks.

## INTRODUCTION

Severe acute respiratory syndrome coronavirus 2 (SARS-CoV-2) infection has affected large numbers of nursing home residents worldwide. The World Health Organization has estimated that as many as half of all coronavirus disease 2019 (COVID-19) deaths in Europe occurred in care homes, and UK Office for National Statistics (ONS) figures confirmed large numbers of deaths in England and Wales in this setting.^1 2^ The true figures are likely to be even higher when indirect mortality effects of the outbreak are accounted for.^3^ Currently, 416,000 people live in UK care homes,^4^ and in addition to their advanced age, the presence of multiple co-morbidities, such as cardiovascular disease and dementia^5^ significantly increases their risk of severe disease and death due to SARS-CoV-2.^6^ In the UK, 89% of deaths due to SARS-CoV-2 are in adults aged >65 years and 43% in those aged >85 years.^7^ Despite the high rates of morbidity and mortality among care home residents, little is known about infection, symptomatology and transmission of SARS-CoV-2 in this highly vulnerable care setting

As with other infections in older people,^8^ early reports have suggested that the clinical manifestations of COVID-19 may be difficult to recognise because the typical symptoms such as cough and breathless may already be present due to other comorbidities,^9^ or they may have non-specific and/or atypical presentations. Despite the use of standard infection control measures, rapid and widespread transmission of SARS-CoV-2 has been reported because of high infectivity both in the pre-symptomatic phase of the illness and a high prevalence of asymptomatic residents in the care home setting.^10^ Within a skilled nursing facility in Washington State USA,^11^ 64% of residents tested positive for SARS-CoV2 and 25% of infected residents died.^10^ Half of those testing positive were asymptomatic at the time of testing, and this was felt to be a key factor contributing to transmission. Traditional approaches to infection control advocated by UK government policy that rely on identification of symptomatic cases and rapid case-isolation may, therefore, be ineffective for limiting the spread of SARS-CoV-2 in care and nursing homes.

Here we report results from an outbreak investigation affecting four UK nursing homes with 394 residents. This investigation was initiated after an unusually high number of residents in one nursing home became unwell in late March/early April 2020. Many residents did not display the typical SARS-CoV-2 symptoms of fever and cough, but died after a short period of becoming acutely unwell.

## METHODS

### Outbreak investigation

On 10 March 2020, when the WHO declared the COVID-19 outbreak a pandemic, there were 373 confirmed cases of COVID-19 in the UK. On 19 March 2020, a resident of Nursing Home A died following an illness consistent with COVID-19. On 25 March, a new resident with confirmed COVID-19 was admitted from home. The first definite de-novo case within Home A was confirmed positive on 26 March. A total of eighteen new residents were admitted to Home A from hospital and 26 residents died between early March and its closure to admissions on 9 April 2020. Because of on-going infections and deaths, comprehensive swabbing of residents and staff was started on 15 April using the increased testing capacity that was put in place. Prior to this date, SARS-CoV-2 testing was being offered for a maximum of five symptomatic residents in each nursing home to confirm an outbreak. Mass testing was possible because of deployment of a high-throughput robotic platform on 10 April, such that local capacity for testing outstripped testing requirement by local hospitals.^12^

The outbreak investigation was convened by the local authority’s Director of Public Health in collaboration with general practitioners, infectious diseases experts, a geriatric clinical outreach team and academics to perform point-prevalence surveys at two time points. Clinical and demographic information was collected alongside comprehensive swabbing of residents. Test results were reported back to residents and care staff promptly for cohorting and implementing additional infection prevention measures where needed. Three phases of testing were performed across the four nursing homes. The second phase involved swabbing of all previously untested or test-negative residents. Those who had tested negative or who were unavailable for testing in the previous round were additionally swabbed one week later.

A convenience sample of asymptomatic staff, chosen to cover the range of roles staff perform within the nursing homes, was also tested to assess infection and potential for transmission. These roles included health care assistants, registered nurses, kitchen staff, administrators, domestic and maintenance staff. Staff testing was conducted in three out of the four homes investigated. Nursing home managers were interviewed to establish staff absence rates before and during the outbreak. Combined oropharyngeal and nasopharyngeal testing with a synthetic fibre swab was used for all the initial tests. The swab was placed into a sterile viral transport medium in keeping with PHE guidance.^13^ Verbal informed consent was sought from those individuals with mental capacity. In residents lacking capacity swabbing was performed in the individual’s best interests based on clinical judgement. Our procedure was revised to bilateral anterior nasal swabbing on 29 April in response to accumulating evidence suggesting equivalent sensitivity of this approach and the better acceptability of this less invasive approach.^14-16^

SARS-CoV-2 was detected using real-time reverse-transcriptase polymerase chain reaction (rt-PCR) in a National Health Service (NHS) diagnostic hospital laboratory using validated assays including the AusDiagnostics, Roche Cobas and Abbott RealTime SARS-CoV-2 assays. If a sample was inconclusive on initial testing, it was repeated using a different assay to confirm a positive/negative result.

### Symptom ascertainment, demographics and comorbidities

Demographic and comorbidity status was obtained from case note review as well as the medical and nursing team. Five key symptoms were recorded including typical (new fever, cough and/or breathlessness) and atypical (newly altered mental status or behaviour, anorexia, diarrhoea or vomiting) features of COVID-19. Retrospective assessment was undertaken for the two weeks prior to the first systematic round of testing, with reassessment performed one week later (mean 6·7 days, SD 2·4) in all those who were asymptomatic at the first testing timepoint.

### Statistical analysis

Data are mainly descriptive. Chi squared test was used to compare categorical data and Wilcoxon rank sum test for non-normally distributed continuous variables. We performed multivariable logistic regression of presenting symptoms in those who had an available test in the initial comprehensive testing round. Analyses were performed in R version 3·6·0.

### Viral Sequencing

To evaluate genomic diversity of the circulating SARS-CoV-2 strains in the care homes, 17 samples were selected from residents (n = 16) and staff (n = 1) who tested positive for SARS-CoV-2. Genomes were extracted, enriched and sequenced following the ARTIC v3 Illumina protocol (see Supplementary Methods). High quality paired-end reads were generated on an Illumina MiSeq instrument and sequences were assembled against the Wuhan-Hu-1-2019 reference genome (NCBI accession MN908947).

### Ethical approval and Patient & Public Involvement

We report the results of an outbreak investigation undertaken as part of usual public health practice for the purposes of defining the cause of the outbreak and how best to manage it. As such the work did not require Research Ethics Committee approval. This is in keeping with UK Health Research Authority guidance and was confirmed with the chair of the West London and GTAC research ethics committee. Patients / members of the public were not involved in its design.

## RESULTS

### Nursing Home Mortality

394 nursing home residents were included in the outbreak investigation. All-cause mortality during the period between 1 March to 1 May 2020 inclusive was 26% (95% CI 22 to 31, N=103). The peak of deaths occurred in the first week of April. A similar time course was observed across the four nursing homes investigated (Figure 1), with marked increases in death rate in homes A, B and D. Comparison with the same time-period in the preceding two years showed a 203% (95% CI 70 to 336) increase in all-cause deaths over the investigation period (Table 1).

**Figure 1.**
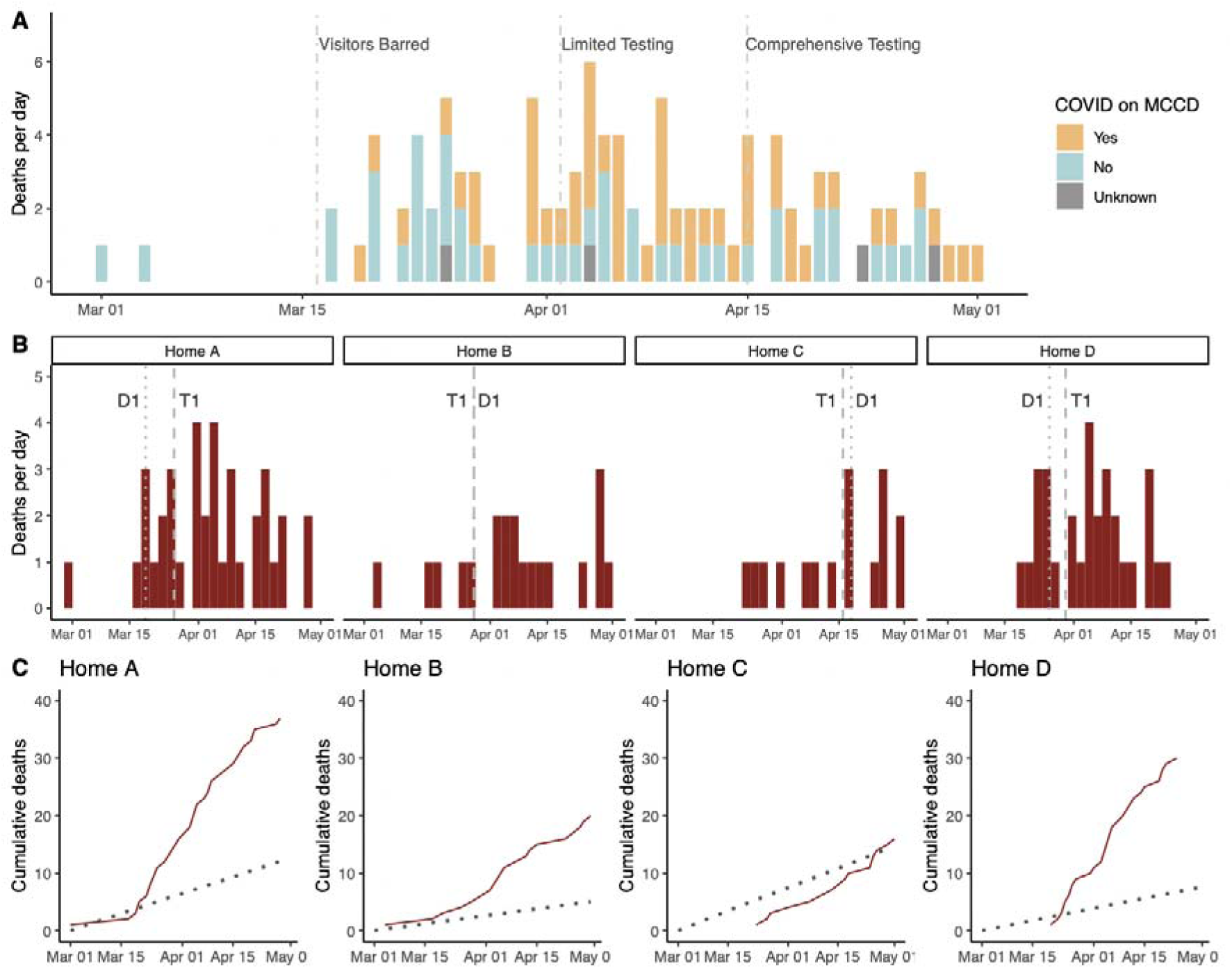
Timeline of COVID-19 outbreak and associated mortality. Panel A – deaths per day throughout the outbreak timeline from 1 March to 1 May 2020 in all four homes, with key dates denoted. Colours relate to the presence of COVID-19 on medical certificate of cause of death (MCCD). Homes were closed to visitors on 16 March; a limited number of tests were made available for symptomatic testing on 2 April by Public Health England; systematic testing within the outbreak investigation commenced on 15 April. Panel B – deaths per day in each nursing home separately. Dates of first positive COVID-19 test (T1) and death-certificate coded COVID-19 death (D1) in each home are shown. Panel C – cumulative number of deaths of all causes, including non-COVID-19, throughout the two-month period (red). Historic average number of deaths throughout the same period in 2018 and 2019 is shown in grey for comparison.

**Table 1.**
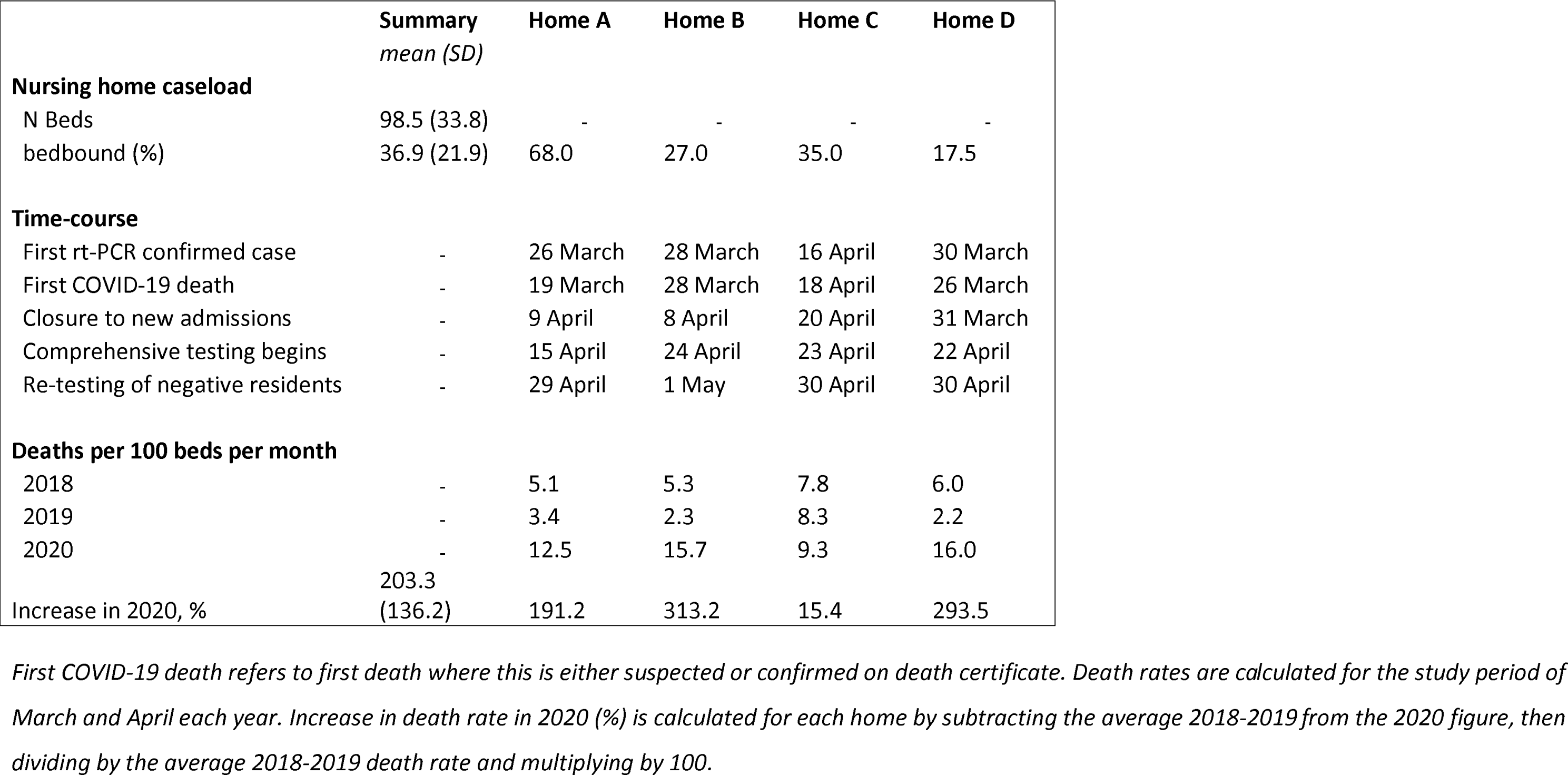
Nursing home information at baseline and during outbreak.

Men had a significantly increased risk of death (48% of deaths vs. 34% in those who survived; whole group 38% male, P=0·020) and there was a trend for the median age to be greater among those who died (P=0·058) (Table 2). Residents had on average three of the co-morbidities assessed (Table 2), with dementia being the most common (57%) followed by cardiovascular disease (CVD; 51%). Cardiovascular disease was the only co-morbidity significantly associated with increased mortality (65% of those who died had CVD versus 45% of survivors; P=0.0010, Figure 2B).

**Table 2.**
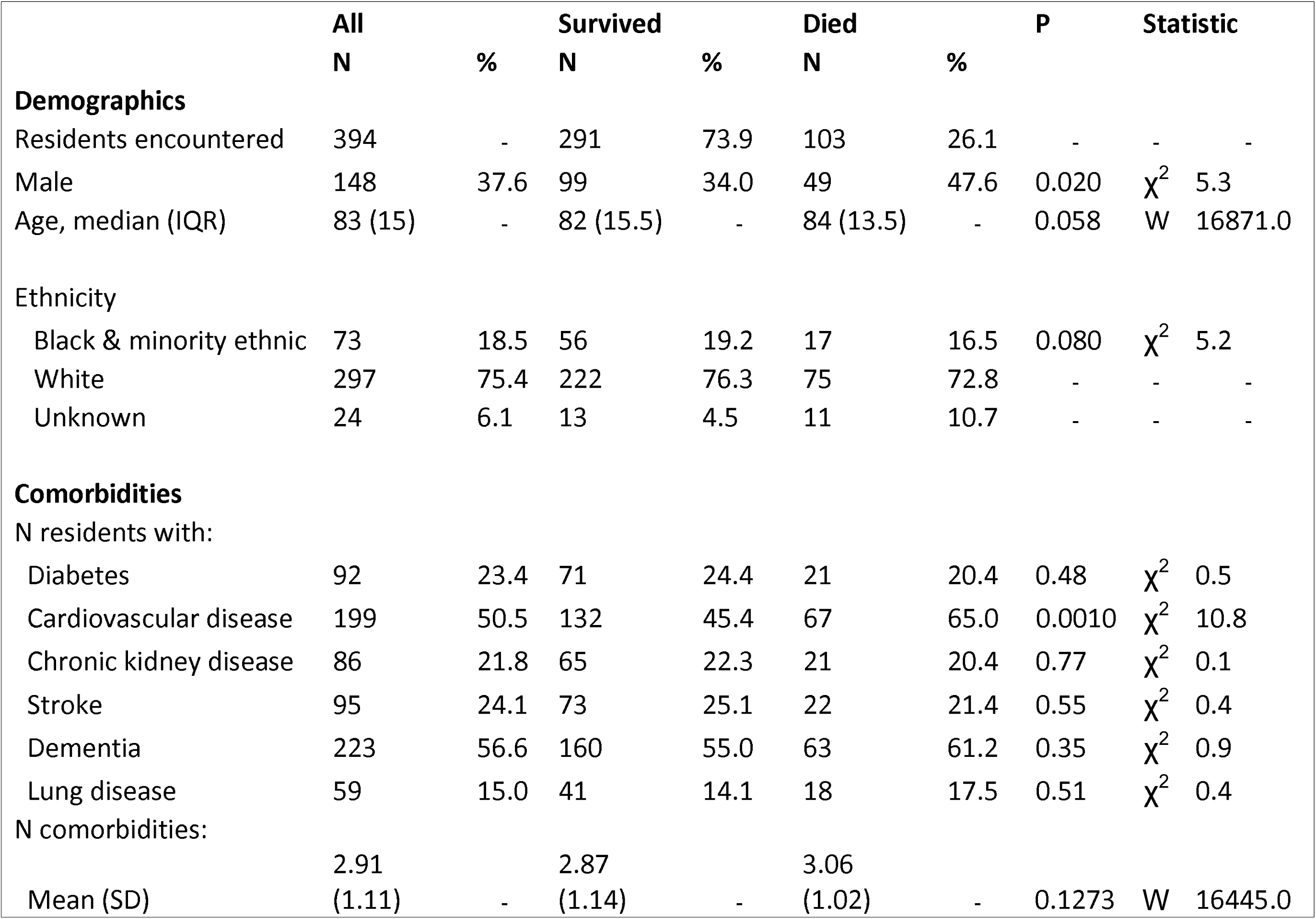
Associations of mortality in nursing homes during COVID-19 outbreak.

**Figure 2.**
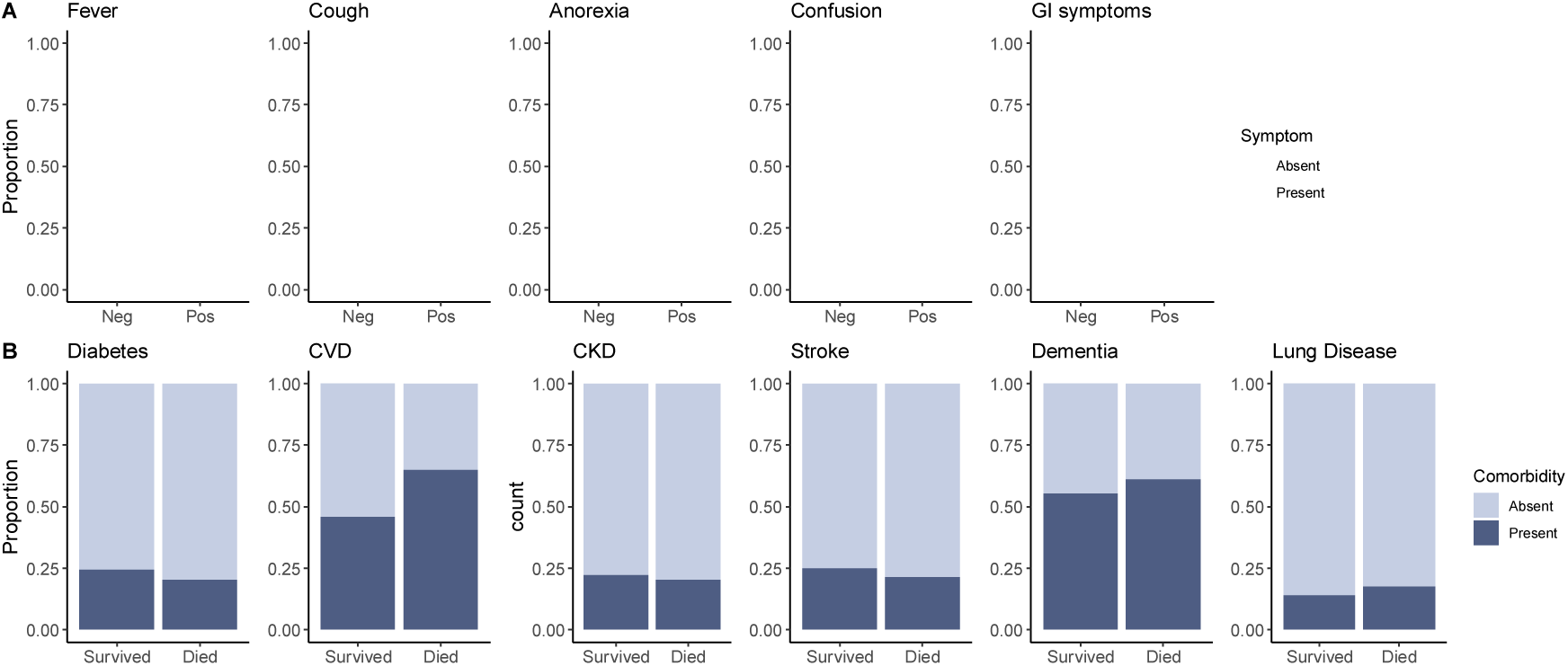
COVID-19 Symptoms, Outcomes and Comorbidities. A – relationship of COVID-19 symptoms in two weeks preceding swabbing with SARS-CoV-2 test result. ‘Confusion’ refers to altered mental status or behaviour. GI symptoms refers to diarrhoea and/or vomiting. B – relationship of comorbidities to all-cause mortality; CVD: cardiovascular disease; CKD: chronic kidney disease; lung disease refers to chronic lung disease.

The medical certificate of cause of death (MCCD) was reviewed for 99/103 residents (Figure 1, panel A). The death certificates listed confirmed or suspected COVID-19 as the underlying cause in the majority of cases (n=53, 54%, 95% CI 44 to 63). The 46 deaths that were coded on the certificate as non-COVID happened earlier in the outbreak on average (mean days since March 1^st^ 33·0, SD 14·2) compared to documented covid-19 deaths (mean 39·1, SD 14·2; W = 1544, P = 0·023). Twelve of these 46 ‘non-covid-19’ deaths were attributed to pneumonia or lower respiratory tract infections and of these three had been tested and were negative for SARS-CoV-2. Sixteen were attributed to frailty or old age, interacting with other comorbidities such as dementia. Two of these residents were rt-PCR positive. In the four individuals where the MCCD was unavailable, the SARS-CoV-2 rt-PCR test was positive and the GP considered the death likely due to COVID-19.

### SARS-CoV-2 Testing Results and Clinical Features

A comprehensive SARS-CoV-2 testing programme for residents was initiated on 15 April 2020. 313 residents were tested in total (Table 3). Fewer than 20 residents declined or were unavailable for testing. Of those tested, 126 (40%, 95% CI 35 to 46) of the residents were positive for SARS-CoV-2. As of 1 May 2020, 21 (17%, 95% CI 10 to 23) residents who tested positive and 8 (4%, 95% CI 1 to 7) with negative tests had died (P=0·0004).

**Table 3.** Clinical symptoms and SARS-CoV-2 rt-PCR test status.

|  | Any result |  | Negative |  | Positive |  | P | Statistic |  |
| --- | --- | --- | --- | --- | --- | --- | --- | --- | --- |
|  | N | % | N | % | N | % |  |  |  |
| **Residents tested** | 313 | - | 187 | 59.7 | 126 | 40.3 | - | - | - |
| **Deaths** | 29 | - | 8 | 4.3 | 21 | 16.7 | 0.0004 | χ<sup>2</sup> | 12.3 |
| **Symptom information** |  |  |  |  |  |  |  |  |  |
| Symptomatic | 109 | 34.8 | 37 | 19.8 | 72 | 57.1 | <0.0001 | χ<sup>2</sup> | 44.7 |
| Asymptomatic | 204 | 65.2 | 150 | 80.2 | 54 | 42.9 | - | - | - |
| N symptoms, mean (SD) | 0.7 (1.1) | - | 0.3 (0.7) | - | 1.2 (1.3) | - | <0.0001 | W | 16542.0 |
| Any typical symptom | 73 | 23.3 | 23 | 12.3 | 50 | 39.7 | <0.0001 | χ<sup>2</sup> | 30.1 |
| Any atypical symptom | 73 | 23.3 | 22 | 11.8 | 51 | 40.5 | <0.0001 | χ<sup>2</sup> | 33.1 |
| Typical symptoms only | 35 | 11.2 | 14 | 7.5 | 21 | 16.7 | 0.019 | χ<sup>2</sup> | 5.5 |
| Atypical symptoms only | 35 | 11.2 | 13 | 7.0 | 22 | 17.5 | 0.0067 | χ<sup>2</sup> | 7.3 |
| **Typical symptoms** |  |  |  |  |  |  |  |  |  |
| Cough / Breathlessness | 57 | 18.2 | 16 | 8.6 | 41 | 32.5 | <0.0001 | χ<sup>2</sup> | 27.5 |
| Fever | 43 | 13.7 | 13 | 7.0 | 30 | 23.8 | <0.0001 | χ<sup>2</sup> | 16.7 |
| **Atypical symptoms** |  |  |  |  |  |  |  |  |  |
| Confusion / Altered behaviour | 62 | 19.8 | 19 | 10.2 | 43 | 34.1 | <0.0001 | χ<sup>2</sup> | 25.7 |
| Anorexia | 44 | 14.1 | 10 | 5.3 | 34 | 27.0 | <0.0001 | χ<sup>2</sup> | 27.4 |
| Diarrhoea / Vomiting | 4 | 1.3 | 2 | 1.1 | 2 | 1.6 | 1.0 | χ<sup>2</sup> | 0.0 |

Residents with negative rt-PCR results who were available for repeat swabbing around 1 week after the initial test, were re-tested (n=173). Combined nasopharyngeal oropharyngeal swabbing was used at the first testing point, whereas anterior nasal swabbing was used for repeat testing. Of these, five (3%, 95% CI 0 to 5) tested positive for SARS-CoV-2 at the second time point.

We assessed the diagnostic value of typical and other COVID-19 symptoms in nursing home residents. 23 (12%, 95% CI 8 to 17) of those who tested negative had displayed one or more of the typical COVID-19 symptoms of cough or fever in the previous two weeks (Table 3). Of 126 (40%) residents who tested positive, well over a third had no symptoms (n=54, 43%, 95% CI 34 to 52). Out of the 72 (57%, 95% CI 49 to 66) residents who did exhibit symptoms, 50 (70% of symptomatic, 95% CI 59 to 80) had any typical symptoms of fever or cough / breathlessness. As many as 22 (31%, 95% CI 20 to 41) of those who were symptomatic with SARS-CoV-2 had none of the typical symptoms.

In our group of residents, the symptom with the strongest independent association with a positive test result for SARS-CoV-2 was new onset anorexia (Figure 2A, Figure 3). Regardless of other symptoms, a resident with new onset anorexia was almost four-times more likely to have SARS-CoV-2 infection than an individual who did not (OR 3·74, 95% CI 1·5 to 9·8). The only other symptoms significantly and independently associated with a positive test result was having either/or cough and shortness of breath (OR 3·72, 95% CI 1·8 to 7·8). Fever was not independently associated with a positive test, neither was altered mental state/behaviour or diarrhoea.

**Figure 3.**
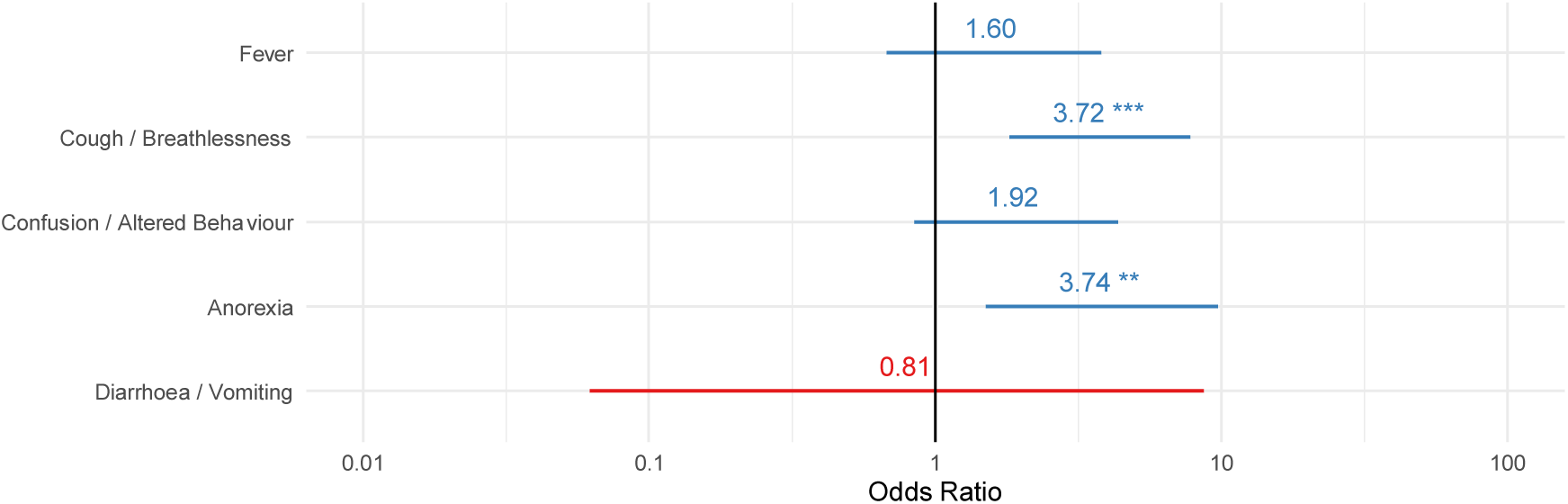
Association of symptoms with a positive SARS-CoV-2 rt-PCR result. Relationship of symptom in preceding two weeks to a positive SARS-CoV-2 result in all residents tested (n=313), displayed as adjusted odds ratios with 95% confidence intervals. Significant predictors in model indicated by ** P<0·01; *** P<0·001.

### Staff sickness and asymptomatic viral carriage

596 members of staff were employed in a variety of roles across the four nursing homes with additional support or illness-cover from agency workers (mean n=149 per home). Staff absence rates due to sickness/self-isolation during the period 1 March to 1 May 2020 were markedly elevated at more than three times the background level (215.9% increase, 95% CI 80 to 352). 70 asymptomatic staff members were tested across three nursing homes (A, C, D). Staff were selected to ensure representation from different roles within the homes. Three (16%, 95% CI -1 to 32) of the 19 staff tested in Home A were positive for SARS-CoV-2; no staff tested positive in Homes C or D (total 4%, 95% CI -1 to 9). Nasopharyngeal/oropharyngeal swabbing was performed in Home A. Anterior nasal swabbing was performed in Homes C and D. (Table 4)

**Table 4.** Nursing home staffing, infection rates and absences.

|  | Summary<br>mean (SD) | Home A | Home B | Home C | Home D |
| --- | --- | --- | --- | --- | --- |
| **Sick or isolating per month** |  |  |  |  |  |
| Baseline, N (%) | - | - | 4.0 (5.1%) | 8.5 (5.7%) | 8.0 (6.7%) |
| Average Mar/Apr 2020, N (%) | - | - | 17.5 (22.1%) | 16.8 (11.2%) | 25.0 (21.0%) |
| Increase in sick/isolating rate (%) | 215.9 (120.0) | - | 337.5 | 97.6 | 212.5 |
| **Asymptomatic testing** |  |  |  |  |  |
| Date(s) of testing | - | April 25 | - | 29 April, 1 May | 1 May |
| Tested, N (% all staff) | 15.6 (11.8%) | 19 (7.6%) | - | 44 (29.3%) | 7 (8.8%) |
| Positive, N (% tested) | 3.0 (4.3%) | 3 (15.7%) | - | 0 (0.0%) | 0 (0.0%) |

### SARS-CoV-2 sequencing

Consensus genomes for 17 positive sample were compared between different residents and different care homes and a Maximum Likelihood phylogenetic tree was calculated (Figure 4). Some SARS-CoV-2 sequence variants were highly similar between residents and/or staff within a single care home. However, this data also showed multiple distinct clusters of SARS-CoV-2 sequence types within single nursing homes.

**Figure 4.**
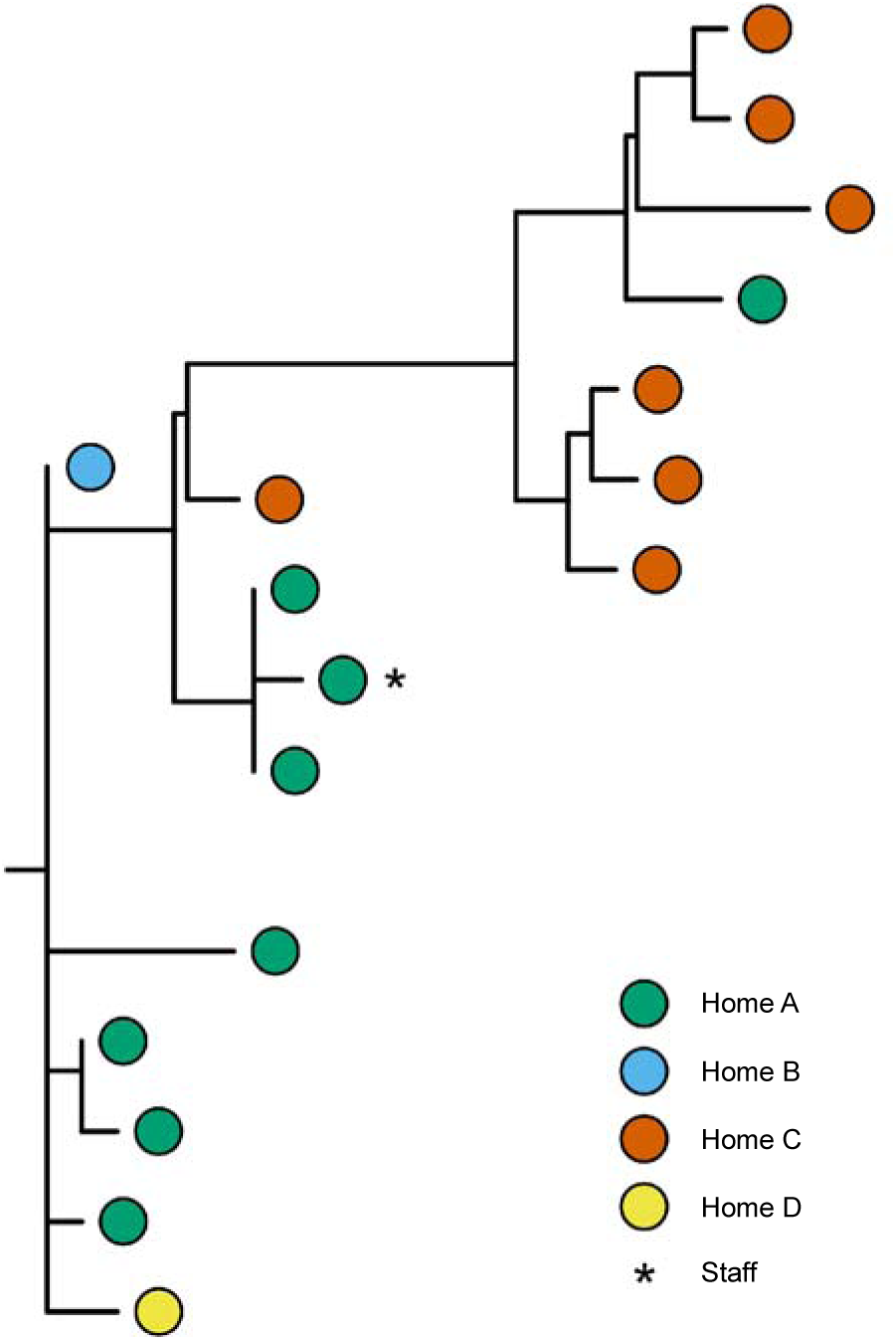
SARS-CoV-2 variants in nursing homes. Maximum Likelihood phylogenetic tree showing SARS-CoV-2 sequence variation across and within the four nursing homes. Coloured dots represent SARS-CoV-2 sequenced from different residents/staff. Horizontal distance in the figure represents number of single nucleotide differences between nodes and branches. Asterisk denotes staff member.

## DISCUSSION

An investigation of COVID-19 outbreaks across four nursing homes in central London identified 103 (26%) fatalities among residents over the two-month period spanning the peak of the London COVID-19 epidemic. These deaths represented a more than two-fold increase compared to the same period in the two previous years, with the increase mostly attributable to three of the four homes investigated. COVID-19 was listed as causative or contributory on just over half of death certificates and was likely underreported in the context of limited testing and non-specific symptoms. Twenty-seven days after the first death and 21 days after the first resident tested positive, we found 126 (40%) of nursing home residents were SARS-CoV-2 positive and 54 (43%) positive residents were asymptomatic. Three of 70 asymptomatic staff (4%) representing various roles in the care home were also positive for SARS-CoV-2. These data catalogue the widespread transmission of SARS-CoV-2 through four UK nursing homes associated with high case fatality rates during the peak pandemic period.

A striking finding of our investigation was that 60% of SARS-CoV-2 positive residents were either asymptomatic or only had atypical symptoms for COVID-19 during the two weeks prior to testing. This was also true of many residents in the days leading up to death indicating that even in severe COVID-19, fever and cough were commonly absent. A novel finding was the strong association between anorexia SARS-CoV-2 positivity in addition to cough/breathlessness. For influenza, atypical symptoms and signs in the elderly and frail are well-recognised, and this is acknowledged in the national guidelines for ‘flu outbreak management.^17^ The initial national drive using symptom-based criteria of fever and cough for testing and isolating individuals may, therefore, have contributed to delays in instituting appropriate infection control measures and, in the nursing home setting, contributed to the large numbers of deaths observed in this highly vulnerable population.

Of the 49 positive residents who were asymptomatic at first testing, only five (10%) went on to develop symptoms. This is in marked contrast to the 89% pre-symptomatic residents identified in the US care home study.^10^ Our first survey occurred later in the outbreak compared to the US study, which may explain the differing pre-symptomatic rates: 21 vs 10 days after the first positive test, and 27 after the first COVID-attributed death. A proportion of our 44 “asymptomatic” residents may have in fact been “post-symptomatic”, with possible symptoms before the two week ascertainment window and unusually prolonged rt-PCR positivity. However, a significant number of the residents are likely to have had true asymptomatic SARS-CoV-2 infection.^18-21^ Serological investigation may help clarify this issue in the future.

We also identified asymptomatic nursing home staff as potential source of viral transmission to the residents. Despite a large proportion of staff self-isolating because they were either unwell or contacts of the confirmed case during the pandemic period, 4% of the working staff who were asymptomatic at the time tested positive for SARS-CoV-2. Additionally, genomic analysis identified one cluster involving one staff member and two residents in the same home. Testing asymptomatic staff members has not been reported in previous studies. This group is likely to have been an important route for SARS-CoV-2 transmission into and within nursing homes. Viral sequencing provided evidence for multiple viral strains within a single nursing home, suggesting that there were multiple introductions into individual nursing homes.

The strengths of this surveillance lie in the large numbers of residents tested across four care homes and over time, with rapid reporting of results and genome sequencing of SARS-CoV-2 isolates to inform transmission patterns. This investigation had some limitations. Firstly, the non-specific nature of COVID-19 symptoms in residents and the lack of availability of tests meant that COVID-19 had to be inferred as the cause of excess deaths in nursing homes prior to our investigation. Since symptoms are often difficult to elicit in residents and are not reliably documented in the records, symptom ascertainment for the 14 days prior to testing was likely to be incomplete. However, we made use of several sources including the residents, care home workers, GPs and records and believe that our data provide a real-life picture. We are only able to infer the role of asymptomatic care staff as potential sources of transmission based on first, staff self-isolation and sickness levels, second, the fact that bedbound residents did not leave the nursing homes and third, similarities in genome sequences of SARS-CoV-2 strains between staff and residents. We could not infer direction of transmission between the two however. A wider exploration of the role of staff was limited by lack of data including reasons for absences, resource issues and lack of compensation for staff members who were aware that if they were SARS-CoV-2 positive, they would have to self-isolate and potentially lose significant income.

Our findings highlight the challenges of controlling SARS-CoV-2 outbreaks in nursing homes. Given the very high mortality observed in the care homes we investigated, this is a pressing issue. During the course of our outbreak investigation, public health policy evolved as the impact of COVID-19 in nursing homes became apparent. At the time of writing, access to testing for all residents and staff has been recommended.^22^ Our results support the need for a policy of universal and systematic testing coupled with a high level of surveillance to ensure the policy is implemented effectively. A regime that involves repeated testing is required because of the potential for asymptomatic residents being infected with SARS-CoV-2 within one week, as we have demonstrated.^14^ Regular testing would enable positive residents and staff to be rapidly identified and appropriate control measures implemented in a timely fashion.

There are, however, practical challenges with more systematic testing of residents and staff in nursing homes. These include logistical barriers to the organisation of mass swabbing and the timely response to results. Nursing homes and linked medical teams will require additional resources to overcome these barriers. Staff testing adds additional challenges including concerns about the impact of staff self-isolation on the ability of a nursing home to continue operating, staff concerns about loss of pay due to self-isolation if positive, and logistical issues such as the need to test temporary staff before allowing them to work in the nursing home.

Research is urgently needed to clarify some key issues to inform infection control guidelines. For example, it is currently unclear how frequently testing should be performed. Longitudinal studies with linked serological assessment are also needed. The most appropriate level of personal protective equipment (PPE) for staff working in nursing homes is still unclear. Basic equipment such as plastic aprons may be inadequate given the close contact often required for bed-bound residents. In addition, it is unclear how best to respond to a positive SARS-CoV-2 test in a nursing home context. There are a range of possible options including cohorting positive residents, restricting seronegative staff from caring for positive residents, enhancing PPE usage during outbreaks and closing the home to visitors and unnecessary personnel earlier during an outbreak. Practical measures to minimise the risk of spread within and between visitors, staff and residents need to be evaluated in a formal and systematic manner. These will need to negotiate the challenges of infection control in nursing homes, including the presence of communal living areas, the wandering behaviours of some residents with dementia, and the constraints of the living environment where decontaminating carpets and soft furnishings may be difficult, and where residents may not easily move rooms.

High mortality rates for nursing homes affected by SARS-CoV-2 necessitate new approaches to infection control. We have shown that the reliance on typical symptoms of COVID-19 infection is inadequate. More comprehensive and regular testing for SARS-CoV-2 is needed at the outset to identify asymptomatic staff and residents, as well as those with atypical symptoms. Further research is needed to inform the most effective and practicable infection control measures to be implemented in an agile way before and during future nursing home outbreaks.

#### Box 1. Research in context

What is already known on this topic

COVID-19 has hit the UK nursing home population hard, with deaths in residential care set to rise above those of all other settings in the UK. Two outbreak investigations from the United States catalogue the rapid spread of COVID-19 in skilled nursing facilities, a potential role for staff in spreading the virus, high fatality rates in residents and a high prevalence of pre-symptomatic infection among residents.^10 11^

What this study adds

We report a detailed investigation of COVID-19 outbreaks in four London nursing homes. We have demonstrated (1) during the earlier part of the epidemic, only a small proportion of deaths among residents were ascribed to COVID-19; death certificate data in the UK nursing home population will likely underestimate the impact of SARS-COV-2; (2) upon systematic testing, 40% of residents tested positive for SARS-CoV-2, many of whom asymptomatic or had atypical symptoms, which means that isolating residents on the basis of cough and/or fever is unlikely to be an effective control measure (3) the identification of asymptomatic staff who tested positive for SARS-CoV-2 highlights this group as potential drivers of transmission within nursing homes. In the absence of effective treatment or immunisation, our data support frequent and repeated screening of all nursing home residents and staff to reduce spread of infection and reduce morbidity and mortality from the disease.

## Data Availability

The data from this public health outbreak investigation are not available for sharing.

## Acknowledgements

The authors would like to thank all those involved in the outbreak investigation particularly the Imperial Care Home Liaison Nurses, Dr Sara Atkin, Dr Tanya Carthy, Dr Sarah Elkin, Dr Anna Wilson, Dr Pandora Wright and Dr Paula Fernandes for their contributions. We would like to thank Kirsten Jensen for help with reagents and Arthi Anand, Vincenzo Pacifico, Katerina Rekopoulou and Paolo Piazza for access to sequencing facilities at Hammersmith Hospital and the Imperial BRC Genomics Facility.

## Author contributions

NL convened the outbreak investigation with FS, DJS, RM and SL. The process was supported by RD, CS, NSNG, RM and CJ who performed testing and clinical/demographic/outcome information ascertainment. PR facilitated laboratory testing. MAC, MS, MP, MC and PSF established the automated RNA extraction platform and MAC, MP carried out RNA extraction from positive samples. LC, MAC, MP, MC and MS performed all the sample preparation for sequencing and analysed the NGS data. LC, MAC, MS and PSF interpreted this data and contributed to the writing of the manuscript. NSNG and CJ performed the analyses. HL assisted analysis and data visualisation. NSNG, CJ, DJS, and FS wrote the draft. AM assisted literature review, and alongside DW and AM writing and revision of the paper. PE and SL critically reviewed and revised the manuscript. All authors contributed to aspects of investigation design/acquisition/analysis/data interpretation; reviewing/revising the paper; approved the final version and agree to be accountable for the work.

## Data sharing

Dissemination of raw data from this investigation is not possible.

## Transparency statement

The lead authors affirm that the manuscript is an honest, accurate, and transparent account of the investigation being reported; that no important aspects of the investigation have been omitted; and that any discrepancies from the study as planned (and, if relevant, registered) have been explained.

## REFERENCES

1. Birnbaum M, Booth W. Nursing homes linked to up to half of coronavirus deaths in Europe, WHO says. Washington, USA. 2020 [updated 23 April. Available from: https://www.washingtonpost.com/world/europe/nursing-homes-coronavirus-deaths-europe/2020/04/23/d635619c-8561-llea-81a3-9690c9881111_story.html accessed 12 May 2020.

2. UK Office for National Statistics. Comparison of weekly death occurrences in England and Wales: up to week ending 24 April 2020: HMSO; 2020 [updated 5 May. Available from: https://www.ons.gov.uk/peoplepopulationandcommunitv/healthandsocialcare/causesofdeath/articles/comparisonofweeklvdeathoccurrencesinenglandandwales/uptoweekending24april2020 accessed 11 May 2020.

3. Comas-Herrera A. Estimates of number of deaths of care home residents linked to the COVID-19 pandemic in England. International Long-Term Care Policy Network: Care Policy and Evaluation Centre, London School of Econonmics, 2020.

4. Competition and Markets Authority. Care homes market study: summary of final report: HMSO; 2017 [updated 30 November. Available from: https://www.gov.uk/government/publications/care-homes-market-studv-summary-of-final-report/care-homes-market-studv-summarv-of-final-report#fn:3 accessed 6th May 2020.

5. Akner G. Analysis of multimorbidity in individual elderly nursing home residents. Development of a multimorbidity matrix. Arch Gerontol Geriatr 2009;49(3):413–9. doi: 10.1016/j.archger.2008.12.009 [published Online First: 2009/02/03]

6. Atkins JL, Masoli JA, Delgado J, et al. PREEXISTING COMORBIDITIES PREDICTING SEVERE COVID-19 IN OLDER ADULTS IN THE UK BIOBANK COMMUNITY COHORT. *medRxiv* 2020:2020.05.06.20092700. doi: 10.1101/2020.05.06.20092700

7. UK Office for National Statistics. Coronavirus (COVID-19) roundup: HMSO; 2020 [updated 5 May. Available from: https://www.ons.gov.uk/peoplepopulationandcommunitv/healthandsocialcare/conditionsanddiseases/articles/coronaviruscovid19roundup/2020-03-26 accessed 6th May 2020.

8. Norman DC. Clinical Features of Infection in Older Adults. Clin Geriatr Med 2016;32(3):433–41. doi: 10.1016/j.cger.2016.02.005 [published Online First: 2016/07/10]

9. British Geriatrics Society. Atypical Covid-19 presentations in older people—the need for continued vigilance 2020 [updated 14 April. Available from: https://www.bgs.org.uk/blog/atypical-covid-19-presentations-in-older-people-the-need-for-continued-vigilance accessed 6 May 2020.

10. Arons MM, Hatfield KM, Reddy SC, et al. Presymptomatic SARS-CoV-2 Infections and Transmission in a Skilled Nursing Facility. N Engl J Med 2020 doi: 10.1056/NEJMoa2008457 [published Online First: 2020/04/25]

11. McMichael TM, Currie DW, Clark S, et al. Epidemiology of Covid-19 in a Long-Term Care Facility in King County, Washington. N Engl J Med 2020 doi: 10.1056/NEJMoa2005412 [published Online First: 2020/03/29]

12. Crone MA, Priestman M, Ciechonska M, et al. A new role for Biofoundries in rapid prototyping, development, and validation of automated clinical diagnostic tests for SARS-CoV-2. *medRxiv* 2020:2020.05.02.20088344. doi: 10.1101/2020.05.02.20088344

13. Public Health England. COVID-19: laboratory investigations and sample requirements for diagnosis: HMSO; 2020 [updated 1 May. Available from: https://www.gov.uk/government/publications/wuhan-novel-coronavirus-guidance-for-clinical-diagnostic-laboratories/laboratorv-investigations-and-sample-requirements-for-diagnosing-and-monitoring-wn-cov-infection accessed 6th May 2020.

14. Wang W, Xu Y, Gao R, et al. Detection of SARS-CoV-2 in Different Types of Clinical Specimens. Jama 2020 doi: 10.1001/jama.2020.3786 [published Online First: 2020/03/12]

15. Kojima N, Turner F, Slepnev V, et al. Self-Collected Oral Fluid and Nasal Swabs Demonstrate Comparable Sensitivity to Clinician Collected Nasopharyngeal Swabs for Covid-19 Detection. *medRxiv* 2020:2020.04.11.20062372. doi: 10.1101/2020.04.11.20062372

16. Tu Y-P, Jennings R, Hart B, et al. Patient-collected tongue, nasal, and mid-turbinate swabs for SARS-CoV-2 yield equivalent sensitivity to health care worker collected nasopharyngeal swabs. *medRxiv* 2020:2020.04.01.20050005. doi: 10.1101/2020.04.01.20050005

17. Public Health England. Infection Prevention and Control: An Outbreak Information Pack for Care Homes. The “Care Home Pack”: HMSO; 2017 [updated October 2019. Available from: https://www.england.nhs.uk/south/wp-content/uploads/sites/6/2019/10/phe-sw-care-home-pack-oct19.pdf accessed 8th May 2020.

18. Lavezzo E, Franchin E, Ciavarella C, et al. Suppression of COVID-19 outbreak in the municipality of Vo, Italy. *medRxiv* 2020:2020.04.17.20053157. doi: 10.1101/2020.04.17.20053157

19. Sutton D, Fuchs K, D’Alton M, et al. Universal Screening for SARS-CoV-2 in Women Admitted for Delivery. New England Journal of Medicine 2020 doi: 10.1056/NEJMc2009316

20. Mizumoto K, Kagaya K, Zarebski A, et al. Estimating the asymptomatic proportion of coronavirus disease 2019 (COVID-19) cases on board the Diamond Princess cruise ship, Yokohama, Japan, 2020. Euro Surveill 2020;25(10) doi: 10.2807/1560-7917.Es.2020.25.10.2000180 [published Online First: 2020/03/19]

21. Nishiura H, Kobayashi T, Miyama T, et al. Estimation of the asymptomatic ratio of novel coronavirus infections (COVID-19). Int J Infect Dis 2020;94:154–55. doi: 10.1016/j.ijid.2020.03.020 [published Online First: 2020/03/18]

22. Hancock M. Health and Social Care Secretary’s statement on coronavirus (COVID-19): 28 April: HMSO; 2020 [Available from: https://www.gov.uk/government/speeches/health-and-social-care-secretarys-statement-on-coronavirus-covid-19-28-april-2020 accessed May 12 2020.

